# COVID-19 outcomes among hospitalized men with or without exposure to alpha-1-adrenergic receptor blocking agents

**DOI:** 10.1101/2021.04.08.21255148

**Authors:** Shilong Li, Tomi Jun, Zichen Wang, Yu-Han Kao, Emilio Schadt, Maximilian F. Konig, Chetan Bettegowda, Joshua T. Vogelstein, Nickolas Papadopoulos, Ramon E. Parsons, Rong Chen, Eric E. Schadt, Li Li, William K. Oh

## Abstract

**Importance:** Alpha-1-adrenergic receptor antagonists (α_1_-blockers) can abrogate pro-inflammatory cytokines and may improve outcomes among patients with respiratory infections. Repurposing readily available drugs such as α_1_-blockers could augment the medical response to the COVID-19 pandemic.

**Objective:** To evaluate the association between α_1_-blocker exposure and COVID-19 mortality

**Design:** Real-world evidence study

**Setting:** Patient level data with 32,355 records tested for SARS-CoV-2 at the Mount Sinai Health System including 8,442 laboratory-confirmed cases extracted from five member hospitals in the New York City metropolitan area.

**Participants:** 2,627 men aged 45 or older admitted with COVID-19 between February 24 and May 31, 2020

**Exposures:** α_1_-blocker use as an outpatient or while admitted for COVID-19

**Main Outcomes and Measures:** In-hospital mortality

**Results:** Men exposed to α_1_-blockers (N=436) were older (median age 73 vs. 64 years, P<0.001) and more likely to have comorbidities than unexposed men (N=2,191). Overall, 758 (28.9%) patients died in hospital, 1,589 (60.5%) were discharged, and 280 (10.7%) were still hospitalized as of May 31, 2020. Outpatient exposure to α_1_-blockers was not associated with COVID-19 hospital outcomes, though there was a trend towards significance (OR 0.749, 95% CI 0.527-1.064; P=0.106). Conversely, inpatient use of α_1_-blockers was independently associated with improved in-hospital mortality in both multivariable logistic (OR 0.633, 95% CI 0.434-0.921; P=0.017) and Cox regression analyses (HR 0.721, 95% CI 0.572-0.908; P=0.006) adjusting for patient demographics, comorbidities, and baseline vitals and labs. Age-stratified analyses suggested greater benefit from inpatient α_1_-blocker use among younger age groups: Age 45-65 OR 0.384, 95% CI 0.164-0.896 (P=0.027); Age 55-75 OR 0.511, 95% CI 0.297-0.880 (P=0.015); Age 65-89 OR 0.810, 95% CI 0.509-1.289 (P=0.374).

**Conclusions and Relevance:** Inpatient α_1_-blocker use was independently associated with improved COVID-19 mortality among hospitalized men. Clinical trials to assess the therapeutic value of α_1_-blockers in COVID-19 are warranted.

## Introduction

Severe coronavirus disease 2019 (COVID-19) has been linked to dysregulated immune responses, including an overexuberant inflammatory response marked by high levels of proinflammatory cytokines such as interleukin-6 (IL-6).^1,2^ Immunosuppressive drugs such as glucocorticoids have become a standard component of treatment for severe COVID-19,^3^ and several trials of anti-cytokine or anti-inflammatory agents are underway or have reported promising results.^4–10^ Despite these advances, there remains a pressing need for safe, effective, and widely available therapeutic options.

Adrenergic signaling has been linked to hyperinflammation in models of bacterial sepsis and cytokine release syndrome. In pre-clinical experiments, a positive feedback loop of adrenergic signaling was identified wherein macrophages responded to catecholamines by producing more catecholamines and inflammatory cytokines; this adrenergic loop could be interrupted by blockade of α_1_-adrenergic receptors with prazosin.^11^ In a retrospective clinical study of patients with acute respiratory distress and pneumonia, exposure to α_1_-adrenergic receptor antagonists (α_1_-blockers) was associated with a significant reduction in risk of mechanical ventilation or death.^12^ Similarly, a recent retrospective analysis of 25,130 patients with COVID-19 across the United States Veterans Health Administration hospital system showed that outpatient exposure to any α_1_-blocker was associated with decreased in-hospital mortality compared to matched controls not on any α_1_-blocker at the time of hospital admission.^13^

These observations have led to the hypothesis that α_1_-blockers in routine clinical use (e.g. prazosin, doxazosin, tamsulosin, etc.) may be repurposed for COVID-19 treatment.^14^ We therefore conducted this real-world evidence study based on electronic medical record (EMR) data to determine whether exposure to α_1_-blockers is independently associated with mortality among patients hospitalized with COVID-19.

## Methods

### Data sources

This retrospective study utilized de-identified electronic medical record (EMR; Epic systems, Verona, WI) data from 5 member hospitals within the Mount Sinai Health System (MSHS) in the New York City metropolitan area. De-identified EMR data were obtained via the Mount Sinai Data Warehouse (https://labs.icahn.mssm.edu/msdw/). We identified 8,442 MSHS patients with PCR confirmed diagnosis of COVID-19 from February 24 through May 31, 2020 during the peak of pandemic in NYC. COVID-19 was diagnosed by real-time reverse transcriptase polymerase chain reaction (RT-PCR)-based clinical tests from nasopharyngeal swab specimens.

We retrieved patient demographics, social history, medication history, and disease comorbidities from the EMR including age, gender, race/ethnicity, smoking status, asthma, chronic obstructive pulmonary disease (COPD), hypertension, obstructive sleep apnea, obesity, diabetes, chronic kidney disease, human immunodeficient virus (HIV) infection, cancer, coronary artery disease, atrial fibrillation, heart failure, chronic viral hepatitis, alcoholic nonalcoholic liver disease, and acute kidney injury (AKI). Patients aged ≥ 89 years were assigned an age of 89 to prevent re-identification. Medications by prescription or hospital administration captured in EMR from January 1, 2019 till May 31, 2020 were included in the medication history. We identified disease comorbidities through their corresponding ICD-10-CM codes before hospital admission and during hospitalization.

We also extracted data from each hospital encounter, including vital sign and laboratory data at the time of presentation, and medications administered during hospitalization. Vital sign and laboratory data extracted included: white blood cell count (WBC), serum creatinine, anion gap, potassium, alanine aminotransferase (ALT), body mass index (BMI), temperature, oxygen saturation, heart rate, respiratory rate, systolic blood pressure (SBP) and diastolic blood pressure (DBP).

We defined three possible outcomes for each hospitalization: in-hospital death (deceased), discharged to home or other locations not associated with acute medical care (recovered), and continued hospitalization (right censored). The duration of hospitalization was calculated from the beginning of the hospital encounter till death or discharge.

This study was approved by the Mount Sinai institutional review board (IRB): IRB-17-01245.

### Study design

This was a retrospective EMR-based study designed to test the independent association of α_1_-blocker exposure with in-hospital death among COVID-19 patients. We first identified 6,218 inpatients positive for COVID-19 in one of 5 hospital systems within the MSHS as of May 31, 2020 (Figure 1). The majority (93%) of α_1_-blocker users in this cohort were men aged 45 or older. We therefore limited the analysis cohort to men aged 45 or older (N=2,627).

**Figure 1.**
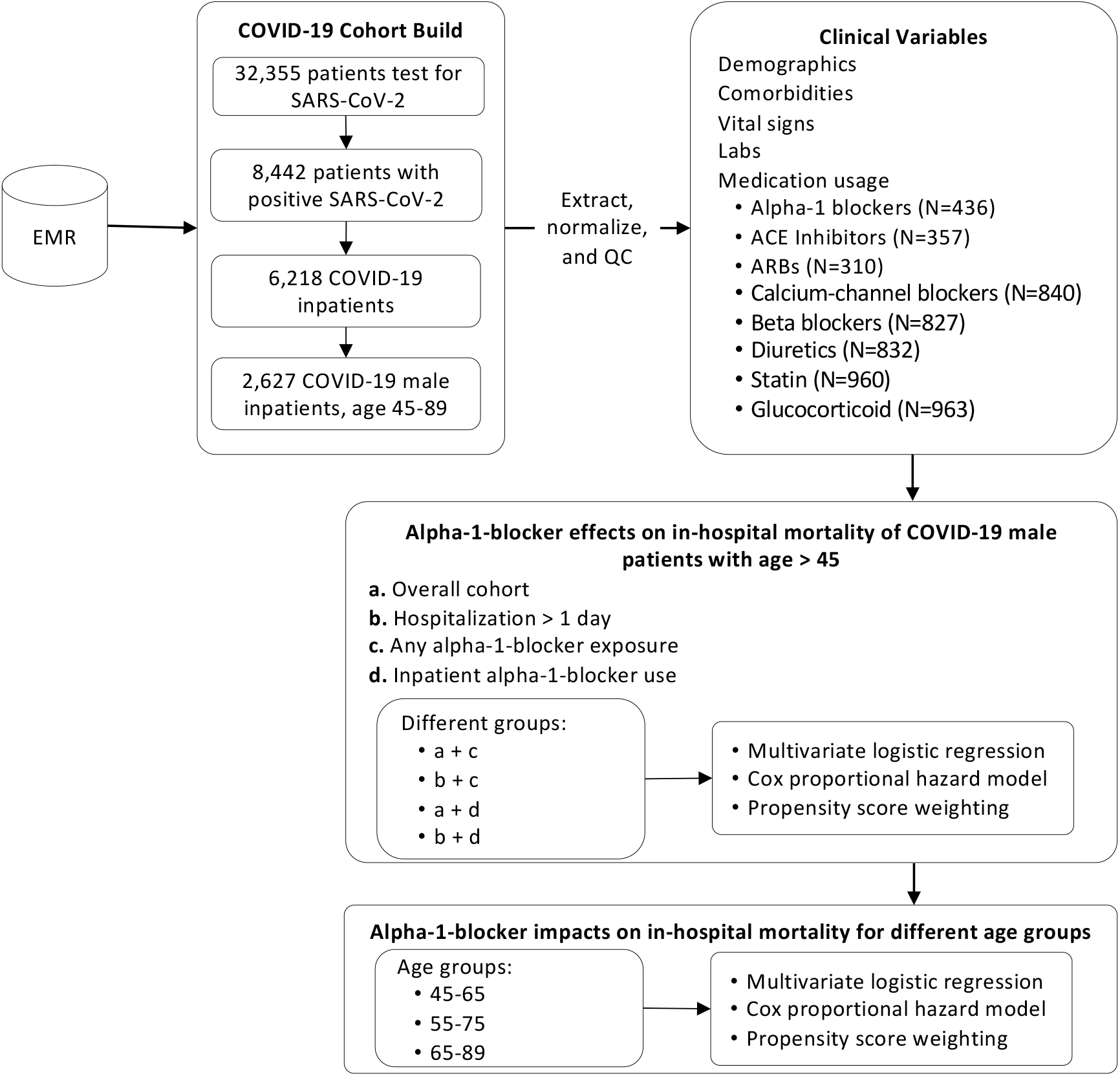
Schematic flowchart of data extraction and analysis plan.

We defined α_1_-blocker exposure as an active prescription from January 1, 2020 for an α_1_-blocker (tamsulosin, alfuzosin, silodosin, terazosin, doxazosin, and prazosin) up to and including hospitalization for COVID-19 (N=436). We further defined a subset of patients (N=343) with documented α_1_-blocker administration during their hospitalization, which we defined as “α_1_-blocker inpatient use”.

Potential confounders in the analysis included demographic characteristics, comorbidities, baseline labs and vitals, and exposure to medications used to treat hypertension, hyperlipidemia, and inflammation. Detailed medication names included in these categories are shown in supplement Table S1.

### Statistical analysis

Patient characteristics were summarized as median and interquartile range (IQR) for continuous variables or mean and standard deviation (SD). We displayed categorical variables as number and percentage (%). We performed a statistical test of hypothesis for differences using the Kruskal-Wallis test or two sample t-test for continuous variables, and the χ^2^ test for categorical variables.

We employed multivariate logistic regression models with potential confounders to estimate the odds ratio and corresponding 95% confidence interval for COVID-19 in-hospital mortality (deceased=1) versus recovery (recovered=0) associated with α_1_-blocker use. We considered the following potential confounders: age, hospital stay duration, race, smoking status, BMI, temperature, O2 saturation, heart rate, respiratory rate, hypertension, asthma, COPD, obstructive sleep apnea, obesity, diabetes mellitus, chronic kidney disease, HIV, cancer, coronary artery disease, atrial fibrillation, heart failure, chronic viral hepatitis, liver disease, AKI, ICU stay, WBC, creatinine, anion gap, potassium, and ALT. We applied the Wald-based power and sample size formulas for logistic regression to calculate the power given the observed sample size.^15^ We used two-tailed test to estimate the probability of event under the null hypothesis which was the in-hospital mortality rate and we used the binomial distribution for α_1_-blocker with the prescription rate as the probability.

To account for right-censored patients (N=280), we used multivariate Cox proportional hazard models to evaluate the association between α_1_-blockers and survival (in-hospital mortality). We estimated the hazard ratio of α_1_-blockers on COVID-19 patient in-hospital survival, adjusting for the same confounders mentioned above.

We utilized propensity score weighting to adjust the potential confounders between those exposed or unexposed to α_1_-blockers. We utilized the R package ‘twang’ and calculated the propensity scores based on average treatment effect on the treated (ATT) and Kolmogorov-Smirnov evaluation criterion. We then employed the ‘survey’ R package by fitting logistic regression models with propensity scores as the weights to assess the α_1_-blocker treatment effect on COVID-19 patients.

Statistical significance was defined as a two-sided P-value < 0.05, unless otherwise noted.

### Sensitivity analysis

To evaluate the robustness of the associations between α_1_-blockers and COVID-19 in-hospital mortality, we performed sensitivity analysis to assess the impact from potential unmeasured or uncontrolled confounding. We calculated E-values for odds and hazard ratios as point estimates, in addition to E-values for their upper confidence limits.^16^ The point estimates represent the strength of unmeasured or uncontrolled confounding required to explain away the observed associations. The upper limits signify the strength of unmeasured or uncontrolled confounding needed to move the confidence interval to include the null, i.e. OR=1 or HR=1. If the effect of unmeasured or uncontrolled confounding is weaker than the calculated E-value, the association of α_1_-blockers with COVID-19 in-hospital mortality could not be fully biased by a weaker confounder.

## Results

### Patient characteristics and outcomes

We identified 6,218 inpatients positive for COVID-19 admitted to one of 5 hospitals within the MSHS as of May 31, 2020. Among these 6,218 patients, 464 patients (7.5%) had been prescribed α_1_-blockers between January 1, 2020 up to and including their hospitalization, with at least 2 months prior to admission; most of these patients were men aged 45 or older (N=436, 93%). Therefore, we limited the analysis cohort to men aged 45 years or older (N=2,627). This cohort was divided into an α_1_-blocker-exposed group (N=436) and an unexposed group (N=2,191). All patients in the α_1_-blocker group had been prescribed α_1_-blockers at some point from January 1, 2020 up to and including their COVID-19 hospitalization (“α_1_-blocker exposure”); a subset of these patients (N=343) had documented α_1_-blocker administration during their hospitalization (“inpatient α_1_-blocker use”).

The α_1_-blocker exposed group was older (median age 73 vs. 64 years, P<0.001) and more likely to have comorbidities than the unexposed group. Chronic diseases such as COPD, hypertension, diabetes mellitus, chronic kidney disease, cancer, and cardiovascular disease were significantly enriched in the α_1_-blocker group. Additional patient demographics for the two groups are shown in Table 1 and Figure S1.

**Table 1.** Baseline characteristics and hospitalization outcomes, by alpha-1-blocker exposure.

|  | Non-Alpha-Blockers | Alpha-Blockers | P-Value |
| --- | --- | --- | --- |
| n | 2191 | 436 |  |
| **Patient Characteristics** |  |  |  |
| Age, median [Q1,Q3] | 64 [56,74] | 73 [66,81] | <0.001 |
| Race, n (%) |  |  | 0.004 |
| African-American | 514 (23.5) | 86 (19.7) |  |
| Asian | 114 (5.2) | 19 (4.4) |  |
| Hispanic | 624 (28.5) | 130 (29.8) |  |
| Other | 336 (15.3) | 49 (11.2) |  |
| Unknown | 69 (3.1) | 10 (2.3) |  |
| White | 534 (24.4) | 142 (32.6) |  |
| BMI (kg/m<sup>2</sup>), median [Q1,Q3] | 27.0 [23.8,30.9] | 26.0 [23.2,30.0] | 0.003 |
| Smoking status, n (%) |  |  | <0.001 |
| Never | 1023 (46.7) | 202 (46.3) |  |
| Not asked | 525 (24.0) | 66 (15.1) |  |
| Quit | 527 (24.1) | 148 (33.9) |  |
| Yes | 116 (5.3) | 20 (4.6) |  |
| Temperature (F), median [Q1,Q3] | 98.6 [97.9,99.8] | 98.5 [98.0,99.5] | 0.365 |
| Max. temperature (F), median [Q1,Q3] | 100.4 [99.1,102.1] | 100.8 [99.6,102.4] | <0.001 |
| O2 saturation (%), median [Q1,Q3] | 96.0 [92.0,98.0] | 95.0 [92.0,97.0] | 0.168 |
| Min. O2 saturation (%), median [Q1,Q3] | 90.0 [82.0,94.0] | 88.0 [78.0,91.0] | <0.001 |
| Heart rate (BPM), median [Q1,Q3] | 94.0 [82.0,108.0] | 91.0 [77.0,104.0] | 0.001 |
| Respiratory rate >25, n (%) | 317 (14.5) | 64 (14.7) | 0.968 |
| High BP (SBP>140 or DBP>90), n (%) | 817 (37.3) | 163 (37.4) | 0.987 |
| **Drugs** |  |  |  |
| ACE inhibitors, n (%) | 267 (12.2) | 90 (20.6) | <0.001 |
| ARBs, n (%) | 222 (10.1) | 88 (20.2) | <0.001 |
| Diuretics, n (%) | 629 (28.7) | 203 (46.6) | <0.001 |
| Beta blockers, n (%) | 623 (28.4) | 204 (46.8) | <0.001 |
| Calcium-channel blockers, n (%) | 627 (28.6) | 213 (48.9) | <0.001 |
| Statin, n (%) | 711 (32.5) | 249 (57.1) | <0.001 |
| Glucocorticoid, n (%) | 745 (34.0) | 218 (50.0) | <0.001 |
| **Comorbidities** |  |  |  |
| Asthma, n (%) | 52 (2.4) | 17 (3.9) | 0.098 |
| COPD, n (%) | 76 (3.5) | 30 (6.9) | 0.002 |
| Hypertension, n (%) | 625 (28.5) | 186 (42.7) | <0.001 |
| Obstructive Sleep Apnea, n (%) | 36 (1.6) | 14 (3.2) | 0.046 |
| Obesity, n (%) | 127 (5.8) | 29 (6.7) | 0.563 |
| Diabetes Mellitus, n (%) | 405 (18.5) | 130 (29.8) | <0.001 |
| Chronic Kidney Disease, n (%) | 216 (9.9) | 75 (17.2) | <0.001 |
| HIV, n (%) | 48 (2.2) | 9 (2.1) | 0.989 |
| Cancer, n (%) | 165 (7.5) | 66 (15.1) | <0.001 |
| Coronary Artery Disease, n (%) | 255 (11.6) | 90 (20.6) | <0.001 |
| Atrial Fibrillation, n (%) | 119 (5.4) | 59 (13.5) | <0.001 |
| Heart Failure, n (%) | 131 (6.0) | 51 (11.7) | <0.001 |
| Chronic Viral Hepatitis, n (%) | 27 (1.2) | 7 (1.6) | 0.691 |
| Liver Disease, n (%) | 47 (2.1) | 13 (3.0) | 0.372 |
| Acute Kidney Injury, n (%) | 150 (6.8) | 47 (10.8) | 0.006 |
| **Labs** |  |  |  |
| WBC (K/uL), median [Q1,Q3] | 7.9 [5.7,10.9] | 7.7 [5.4,11.4] | 0.531 |
| Creatinine > 1.2, n (%) | 903 (41.2) | 231 (53.0) | <0.001 |
| Anion Gap (mEq/L), median [Q1,Q3] | 12.3 [10.7,15.0] | 12.4 [10.2,15.0] | 0.413 |
| Potassium (mmol/L), median [Q1,Q3] | 4.4 [4.0,4.8] | 4.3 [3.9,4.8] | 0.622 |
| ALT (U/L), median [Q1,Q3] | 34.0 [21.0,57.0] | 29.0 [18.0,50.0] | <0.001 |
| **Hospital Conditions** |  |  |  |
| ICU, n (%) | 431 (19.7) | 112 (25.7) | 0.006 |
| Days after infection, median [Q1,Q3] | 4.7 [1.1,9.5] | 7.0 [3.6,11.5] | <0.001 |
| Hospital status, n (%) |  |  | <0.001 |
| Censored | 214 (9.8) | 66 (15.1) |  |
| Deceased | 617 (28.2) | 141 (32.3) |  |
| Discharged | 1360 (62.1) | 229 (52.5) |  |

Abbreviations: BMI, body mass index; BPM, beats per minute; BP, blood pressure; SBP, systolic blood pressure; DBP, diastolic blood pressure; ACE, angiotensin-converting enzyme; ARBs, angiotensin II receptor blockers; COPD, chronic obstructive pulmonary disease; HIV, human immunodeficient virus; WBC, white blood cell count; ALT, alanine aminotransferase

Overall, 758 (28.9%) patients died, 1,589 (60.5%) were discharged, and 280 (10.7%) were still hospitalized as of May 31, 2020. Unadjusted in-hospital mortality was 32.3% in the exposed group and 28.2% in the unexposed group.

### α_1_-blocker exposure and COVID-19 in-hospital mortality

We evaluated the association of α_1_-blockers on COVID-19 patients with known outcomes (death: N=758; discharge: N=1,589) using multivariate logistic-regression models. In the overall population, α_1_-blocker exposure was not significantly associated with in-hospital mortality (OR 0.749; 95% CI, 0.527-1.064; P=0.106) with the power β = 0.62 based on the sample size of 2347. There was a trend towards a protective association for α_1_-blocker exposure in the subset of patients who were hospitalized for at least 24 hours, though this was not significant (N=2,276; OR 0.722; 95% CI, 0.505-1.034; P=0.075) (Table 2). This might be due to limited sample size (β = 0.71 in the Wald-based power analysis).

**Table 2.** Multivariable logistic regression results for COVID-19 in-hospital mortality, by cohort (overall vs. hospitalization >1 day) and alpha-1-blocker exposure (any vs. inpatient use)

|  | Overall cohort, any exposure<br>(N=2347) |  | Overall cohort, inpatient use<br>(N=2276) |  | Hospital stay > 1 day, any<br>exposure (N=1789) |  | Hospital stay > 1 day,<br>inpatient use (N=1744) |  |
| --- | --- | --- | --- | --- | --- | --- | --- | --- |
|  | Adjusted OR (95%<br>CI) | p-value | Adjusted OR (95%<br>CI) | p-value | Adjusted OR<br>(95% CI) | p-value | Adjusted OR<br>(95% CI) | p-value |
| **Drugs** |  |  |  |  |  |  |  |  |
| Alpha-1 blockers | 0.749 (0.527-1.064) | 0.106 | 0.633 (0.434-0.921) | 0.017 | 0.722 (0.505-1.034) | 0.075 | 0.607 (0.415-0.890) | 0.010 |
| ACE inhibitors | 1.331 (0.880-2.014) | 0.175 | 1.237 (0.808-1.893) | 0.327 | 1.317 (0.861-2.016) | 0.205 | 1.220 (0.787-1.891) | 0.373 |
| ARBs | 1.005 (0.660-1.528) | 0.983 | 1.030 (0.667-1.592) | 0.893 | 0.872 (0.562-1.353) | 0.541 | 0.900 (0.572-1.419) | 0.652 |
| Diuretics | 1.595 (1.147-2.217) | 0.006 | 1.571 (1.123-2.199) | 0.008 | 1.520 (1.084-2.132) | 0.015 | 1.464 (1.037-2.067) | 0.030 |
| Beta blockers | 1.448 (1.067-1.966) | 0.017 | 1.534 (1.123-2.094) | 0.007 | 1.505 (1.103-2.054) | 0.010 | 1.578 (1.148-2.168) | 0.005 |
| Calcium-channel<br>blockers | 0.736 (0.535-1.012) | 0.059 | 0.698 (0.503-0.968) | 0.031 | 0.758 (0.547-1.050) | 0.095 | 0.710 (0.508-0.991) | 0.044 |
| Statin | 0.866 (0.624-1.200) | 0.386 | 0.903 (0.647-1.261) | 0.550 | 0.925 (0.661-1.293) | 0.646 | 0.967 (0.686-1.361) | 0.844 |
| Glucocorticoid | 1.368 (0.981-1.904) | 0.064 | 1.422 (1.014-1.994) | 0.041 | 1.467 (1.044-2.061) | 0.027 | 1.495 (1.058-2.115) | 0.023 |
| **Patient Characteristics** |  |  |  |  |  |  |  |  |
| Age | 1.079 (1.063-1.095) | <0.001 | 1.081 (1.064-1.099) | <0.001 | 1.084 (1.067-1.102) | <0.001 | 1.085 (1.068-1.103) | <0.001 |
| Race: African American | 0.701 (0.463-1.063) | 0.095 | 0.722 (0.473-1.102) | 0.131 | 0.826 (0.539-1.266) | 0.381 | 0.838 (0.542-1.293) | 0.424 |
| Race: Asian | 0.637 (0.311-1.307) | 0.219 | 0.554 (0.260-1.179) | 0.126 | 0.708 (0.342-1.468) | 0.354 | 0.600 (0.279-1.294) | 0.193 |
| Race: Hispanic | 0.776 (0.525-1.147) | 0.202 | 0.786 (0.526-1.174) | 0.238 | 0.872 (0.582-1.306) | 0.507 | 0.872 (0.577-1.319) | 0.516 |
| Race: Other | 0.650 (0.405-1.043) | 0.074 | 0.673 (0.417-1.087) | 0.105 | 0.763 (0.470-1.237) | 0.273 | 0.780 (0.479-1.273) | 0.321 |
| Race: Unknown | 0.725 (0.293-1.795) | 0.488 | 0.737 (0.296-1.839) | 0.513 | 0.818 (0.329-2.030) | 0.664 | 0.825 (0.330-2.065) | 0.682 |
| Smoking Status: Not<br>Asked | 1.001 (0.678-1.477) | 0.996 | 0.982 (0.662-1.456) | 0.930 | 0.932 (0.626-1.388) | 0.731 | 0.941 (0.629-1.406) | 0.765 |
| Smoking Status: Quit | 1.351 (0.949-1.923) | 0.095 | 1.302 (0.906-1.872) | 0.154 | 1.306 (0.909-1.876) | 0.148 | 1.309 (0.902-1.896) | 0.156 |
| Smoking Status: Yes | 1.274 (0.637-2.545) | 0.494 | 1.353 (0.676-2.710) | 0.393 | 1.232 (0.604-2.514) | 0.566 | 1.335 (0.652-2.732) | 0.430 |
| BMI | 1.011 (0.989-1.035) | 0.326 | 1.013 (0.991-1.037) | 0.251 | 1.013 (0.990-1.038) | 0.271 | 1.014 (0.991-1.039) | 0.233 |
| Temperature | 0.991 (0.930-1.057) | 0.779 | 0.993 (0.931-1.061) | 0.842 | 0.991 (0.928-1.059) | 0.781 | 0.992 (0.928-1.061) | 0.820 |
| Max. Temperature | 1.221 (1.122-1.331) | <0.001 | 1.212 (1.112-1.322) | <0.001 | 1.262 (1.154-1.380) | <0.001 | 1.247 (1.139-1.366) | <0.001 |
| O2 Saturation | 1.013 (0.991-1.037) | 0.243 | 1.014 (0.992-1.037) | 0.220 | 1.006 (0.983-1.029) | 0.617 | 1.008 (0.984-1.031) | 0.513 |
| Min. O2 Saturation | 0.932 (0.919-0.946) | <0.001 | 0.932 (0.919-0.946) | <0.001 | 0.934 (0.920-0.947) | <0.001 | 0.933 (0.919-0.946) | <0.001 |
| Heart Rate | 0.996 (0.989-1.004) | 0.353 | 0.995 (0.987-1.003) | 0.201 | 0.995 (0.987-1.003) | 0.219 | 0.994 (0.986-1.002) | 0.164 |
| Respiratory Rate > 25 | 1.699 (1.151-2.504) | 0.007 | 1.649 (1.107-2.452) | 0.014 | 1.511 (1.010-2.261) | 0.045 | 1.476 (0.977-2.228) | 0.064 |
| High BP (SBP>140 or<br>DBP>90) | 1.161 (0.863-1.560) | 0.324 | 1.169 (0.863-1.584) | 0.312 | 1.219 (0.900-1.650) | 0.200 | 1.234 (0.905-1.682) | 0.184 |
| **Comorbidities** |  |  |  |  |  |  |  |  |
| Asthma | 0.259 (0.088-0.765) | 0.015 | 0.299 (0.099-0.900) | 0.032 | 0.280 (0.095-0.823) | 0.021 | 0.323 (0.109-0.962) | 0.042 |
| COPD | 1.505 (0.771-2.939) | 0.231 | 1.342 (0.672-2.678) | 0.405 | 1.498 (0.752-2.980) | 0.250 | 1.296 (0.633-2.651) | 0.479 |
| Hypertension | 0.825 (0.570-1.196) | 0.311 | 0.851 (0.580-1.249) | 0.409 | 0.817 (0.558-1.196) | 0.299 | 0.837 (0.564-1.241) | 0.376 |
| Obstructive Sleep Apnea | 3.025 (1.175-7.791) | 0.022 | 3.019 (1.163-7.838) | 0.023 | 3.074 (1.164-8.125) | 0.023 | 3.152 (1.179-8.415) | 0.022 |
| Obesity | 1.048 (0.568-1.935) | 0.880 | 1.064 (0.567-1.998) | 0.847 | 0.919 (0.485-1.742) | 0.797 | 0.924 (0.479-1.784) | 0.815 |
| Diabetes Mellitus | 1.043 (0.710-1.533) | 0.829 | 0.986 (0.664-1.465) | 0.946 | 1.099 (0.737-1.639) | 0.643 | 1.025 (0.679-1.547) | 0.907 |
| Chronic Kidney Disease | 1.188 (0.737-1.914) | 0.479 | 1.264 (0.774-2.063) | 0.349 | 1.322 (0.810-2.158) | 0.264 | 1.412 (0.854-2.333) | 0.179 |
| HIV | 1.846 (0.688-4.948) | 0.224 | 1.696 (0.600-4.792) | 0.319 | 1.702 (0.604-4.802) | 0.314 | 1.480 (0.494-4.433) | 0.484 |
| Cancer | 0.959 (0.601-1.530) | 0.861 | 0.862 (0.530-1.398) | 0.546 | 0.882 (0.546-1.426) | 0.609 | 0.831 (0.506-1.365) | 0.464 |
| Coronary Artery Disease | 0.807 (0.514-1.265) | 0.350 | 0.783 (0.492-1.249) | 0.305 | 0.880 (0.550-1.406) | 0.593 | 0.899 (0.555-1.455) | 0.663 |
| Atrial Fibrillation | 0.779 (0.457-1.330) | 0.360 | 0.740 (0.428-1.279) | 0.281 | 0.768 (0.444-1.328) | 0.345 | 0.726 (0.414-1.275) | 0.266 |
| Heart Failure | 1.004 (0.576-1.749) | 0.989 | 1.041 (0.587-1.846) | 0.891 | 0.838 (0.466-1.507) | 0.554 | 0.836 (0.454-1.540) | 0.566 |
| Chronic Viral Hepatitis | 0.546 (0.146-2.040) | 0.368 | 0.596 (0.158-2.255) | 0.446 | 0.580 (0.155-2.179) | 0.420 | 0.630 (0.166-2.394) | 0.497 |
| Liver Disease | 1.448 (0.593-3.536) | 0.417 | 1.486 (0.607-3.636) | 0.386 | 1.573 (0.639-3.873) | 0.325 | 1.613 (0.654-3.983) | 0.299 |
| Acute Kidney Injury | 1.157 (0.689-1.944) | 0.581 | 1.009 (0.582-1.747) | 0.975 | 1.204 (0.708-2.048) | 0.493 | 1.107 (0.635-1.933) | 0.718 |
| **Labs** |  |  |  |  |  |  |  |  |
| WBC | 1.028 (1.000-1.058) | 0.052 | 1.029 (1.001-1.060) | 0.043 | 1.019 (0.988-1.052) | 0.234 | 1.020 (0.988-1.053) | 0.212 |
| Creatinine > 1.2 | 1.660 (1.184-2.330) | 0.003 | 1.598 (1.130-2.261) | 0.008 | 1.462 (1.035-2.067) | 0.031 | 1.438 (1.009-2.048) | 0.045 |
| Anion Gap | 1.074 (1.035-1.114) | <0.001 | 1.075 (1.036-1.116) | <0.001 | 1.063 (1.024-1.103) | 0.001 | 1.063 (1.023-1.104) | 0.001 |
| Potassium | 1.223 (1.016-1.471) | 0.033 | 1.191 (0.985-1.441) | 0.072 | 1.219 (1.010-1.473) | 0.039 | 1.184 (0.975-1.438) | 0.088 |
| ALT | 1.001 (0.999-1.003) | 0.309 | 1.001 (0.999-1.004) | 0.271 | 1.001 (0.999-1.003) | 0.349 | 1.001 (0.999-1.003) | 0.364 |
| **Hospital Conditions** |  |  |  |  |  |  |  |  |
| ICU | 3.547 (2.433-5.165) | <0.001 | 3.740 (2.550-5.479) | <0.001 | 3.219 (2.197-4.711) | <0.001 | 3.425 (2.323-5.048) | <0.001 |
| Duration (days) | 0.956 (0.933-0.979) | <0.001 | 0.958 (0.934-0.981) | <0.001 | 0.959 (0.936-0.983) | 0.001 | 0.962 (0.939-0.986) | 0.002 |

Abbreviations: ACE, angiotensin-converting enzyme; ARBs, angiotensin II receptor blockers; BMI, body mass index; BP, blood pressure; SBP, systolic blood pressure; DBP, diastolic blood pressure; COPD, chronic obstructive pulmonary disease; HIV, human immunodeficient virus; WBC, white blood cell count; ALT, alanine aminotransferase

To account for right-censored patients (N=280), we used Cox proportional hazard models to evaluate the association of α_1_-blocker exposure and COVID-19 in-hospital survival. There was a trend towards a protective effect of α_1_-blocker exposure in the overall cohort (HR 0.808; 95% CI, 0.652-1.002; P=0.052) and among those hospitalized for at least 24 hours (HR 0.823; 95% CI, 0.660-1.027; P=0.085), though these associations were not significant (Table 3).

**Table 3.** Multivariable Cox regression results for COVID-19 in-hospital mortality, by cohort (overall vs. hospitalization >1 day) and alpha-1-blocker exposure (any vs. inpatient use)

|  | Overall cohort, any exposure<br>(N=2627) |  | Overall cohort, inpatient use<br>(N=2534) |  | Hospital stay > 1 day, any<br>exposure (N=2037) |  | Hospital stay > 1 day,<br>inpatient use (N=1973) |  |
| --- | --- | --- | --- | --- | --- | --- | --- | --- |
|  | HR (95% CI) | P-value | HR (95% CI) | P-value | HR (95% CI) | P-value | HR (95% CI) | P-value |
| **Drugs** |  |  |  |  |  |  |  |  |
| Alpha-1 blockers | 0.808 (0.652-1.002) | 0.052 | 0.721 (0.572-0.908) | 0.006 | 0.823 (0.660-1.027) | 0.085 | 0.751 (0.593-0.950) | 0.017 |
| ACE inhibitors | 1.166 (0.910-1.494) | 0.224 | 1.149 (0.887-1.487) | 0.292 | 1.151 (0.892-1.486) | 0.280 | 1.121 (0.859-1.462) | 0.402 |
| ARBs | 1.330 (1.035-1.709) | 0.026 | 1.319 (1.018-1.710) | 0.036 | 1.152 (0.880-1.509) | 0.302 | 1.145 (0.867-1.512) | 0.341 |
| Diuretics | 0.895 (0.729-1.098) | 0.286 | 0.863 (0.700-1.065) | 0.169 | 0.859 (0.694-1.064) | 0.164 | 0.825 (0.663-1.026) | 0.084 |
| Beta blockers | 1.073 (0.895-1.287) | 0.446 | 1.090 (0.905-1.313) | 0.363 | 1.100 (0.912-1.327) | 0.317 | 1.120 (0.925-1.356) | 0.247 |
| Calcium-channel blockers | 0.637 (0.530-0.766) | <0.001 | 0.613 (0.508-0.740) | <0.001 | 0.649 (0.537-0.785) | <0.001 | 0.629 (0.518-0.764) | <0.001 |
| Statin | 0.970 (0.800-1.175) | 0.753 | 0.951 (0.778-1.162) | 0.621 | 1.007 (0.825-1.228) | 0.949 | 0.999 (0.812-1.228) | 0.990 |
| Glucocorticoid | 0.767 (0.627-0.938) | 0.010 | 0.789 (0.642-0.971) | 0.025 | 0.801 (0.649-0.988) | 0.038 | 0.820 (0.661-1.016) | 0.070 |
| **Patient Characteristics** |  |  |  |  |  |  |  |  |
| Age | 1.035 (1.025-1.044) | <0.001 | 1.033 (1.024-1.043) | <0.001 | 1.037 (1.027-1.047) | <0.001 | 1.035 (1.025-1.045) | <0.001 |
| Race: African American | 0.813 (0.633-1.043) | 0.104 | 0.824 (0.639-1.064) | 0.137 | 0.906 (0.699-1.175) | 0.457 | 0.909 (0.698-1.184) | 0.480 |
| Race: Asian | 0.829 (0.536-1.284) | 0.401 | 0.773 (0.488-1.224) | 0.272 | 0.841 (0.533-1.326) | 0.456 | 0.760 (0.470-1.230) | 0.264 |
| Race: Hispanic | 0.881 (0.698-1.112) | 0.286 | 0.866 (0.682-1.099) | 0.237 | 0.928 (0.727-1.184) | 0.548 | 0.919 (0.716-1.178) | 0.504 |
| Race: Other | 0.972 (0.738-1.279) | 0.839 | 0.972 (0.737-1.284) | 0.843 | 1.043 (0.784-1.389) | 0.773 | 1.035 (0.775-1.382) | 0.816 |
| Race: Unknown | 0.961 (0.578-1.599) | 0.879 | 0.982 (0.589-1.636) | 0.944 | 1.043 (0.625-1.741) | 0.871 | 1.060 (0.634-1.772) | 0.824 |
| Smoking Status: Not Asked | 1.089 (0.865-1.371) | 0.467 | 1.084 (0.858-1.369) | 0.499 | 1.065 (0.840-1.351) | 0.603 | 1.085 (0.853-1.379) | 0.507 |
| Smoking Status: Quit | 1.262 (1.022-1.558) | 0.030 | 1.277 (1.027-1.587) | 0.028 | 1.251 (1.006-1.558) | 0.044 | 1.293 (1.033-1.620) | 0.025 |
| Smoking Status: Yes | 1.090 (0.714-1.664) | 0.690 | 1.104 (0.719-1.695) | 0.652 | 1.065 (0.689-1.645) | 0.778 | 1.094 (0.704-1.702) | 0.689 |
| BMI | 1.008 (0.996-1.020) | 0.168 | 1.008 (0.996-1.021) | 0.172 | 1.009 (0.997-1.022) | 0.134 | 1.009 (0.997-1.021) | 0.153 |
| Temperature | 1.021 (0.972-1.072) | 0.406 | 1.013 (0.966-1.062) | 0.595 | 1.012 (0.964-1.062) | 0.621 | 1.006 (0.960-1.055) | 0.789 |
| Max. Temperature | 1.016 (0.963-1.071) | 0.562 | 1.013 (0.959-1.069) | 0.646 | 1.061 (1.007-1.117) | 0.025 | 1.058 (1.004-1.115) | 0.035 |
| O2 Saturation | 0.990 (0.979-1.000) | 0.057 | 0.991 (0.980-1.002) | 0.112 | 0.988 (0.977-0.999) | 0.036 | 0.989 (0.978-1.001) | 0.069 |
| Min. O2 Saturation | 0.991 (0.988-0.995) | <0.001 | 0.991 (0.988-0.995) | <0.001 | 0.993 (0.989-0.997) | 0.001 | 0.993 (0.989-0.997) | <0.001 |
| Heart Rate | 1.000 (0.995-1.004) | 0.939 | 0.999 (0.994-1.003) | 0.605 | 1.000 (0.995-1.005) | 0.920 | 0.999 (0.994-1.004) | 0.779 |
| Respiratory Rate > 25 | 1.155 (0.939-1.420) | 0.173 | 1.136 (0.918-1.405) | 0.241 | 1.109 (0.893-1.378) | 0.349 | 1.101 (0.882-1.374) | 0.395 |
| High BP (SBP>140 or DBP>90) | 1.070 (0.898-1.275) | 0.451 | 1.095 (0.915-1.310) | 0.323 | 1.135 (0.946-1.361) | 0.172 | 1.156 (0.960-1.392) | 0.125 |
| **Comorbidities** |  |  |  |  |  |  |  |  |
| Asthma | 0.501 (0.234-1.072) | 0.075 | 0.534 (0.249-1.147) | 0.108 | 0.536 (0.250-1.149) | 0.109 | 0.582 (0.271-1.250) | 0.165 |
| COPD | 1.254 (0.871-1.805) | 0.224 | 1.121 (0.758-1.657) | 0.568 | 1.280 (0.876-1.872) | 0.202 | 1.130 (0.752-1.699) | 0.557 |
| Hypertension | 0.877 (0.696-1.103) | 0.261 | 0.881 (0.695-1.118) | 0.298 | 0.857 (0.676-1.088) | 0.205 | 0.850 (0.665-1.086) | 0.193 |
| Obstructive Sleep Apnea | 1.257 (0.721-2.193) | 0.420 | 1.405 (0.794-2.486) | 0.242 | 1.138 (0.631-2.054) | 0.667 | 1.267 (0.690-2.329) | 0.445 |
| Obesity | 0.954 (0.667-1.363) | 0.795 | 0.998 (0.695-1.434) | 0.992 | 0.954 (0.658-1.382) | 0.802 | 0.999 (0.685-1.456) | 0.996 |
| Diabetes Mellitus | 0.986 (0.785-1.239) | 0.904 | 0.981 (0.776-1.240) | 0.873 | 0.970 (0.765-1.229) | 0.798 | 0.954 (0.748-1.216) | 0.703 |
| Chronic Kidney Disease | 0.920 (0.694-1.220) | 0.564 | 0.958 (0.718-1.278) | 0.769 | 1.050 (0.784-1.405) | 0.743 | 1.090 (0.809-1.468) | 0.572 |
| HIV | 0.943 (0.515-1.728) | 0.850 | 0.878 (0.467-1.653) | 0.688 | 0.901 (0.478-1.698) | 0.748 | 0.848 (0.437-1.644) | 0.625 |
| Cancer | 1.255 (0.947-1.663) | 0.114 | 1.213 (0.903-1.627) | 0.199 | 1.237 (0.922-1.660) | 0.156 | 1.200 (0.884-1.630) | 0.243 |
| Coronary Artery Disease | 1.190 (0.917-1.544) | 0.192 | 1.231 (0.940-1.613) | 0.131 | 1.236 (0.941-1.625) | 0.128 | 1.298 (0.978-1.721) | 0.071 |
| Atrial Fibrillation | 1.306 (0.944-1.807) | 0.107 | 1.353 (0.966-1.893) | 0.078 | 1.278 (0.905-1.806) | 0.163 | 1.295 (0.904-1.854) | 0.158 |
| Heart Failure | 1.499 (1.092-2.058) | 0.012 | 1.538 (1.108-2.135) | 0.010 | 1.314 (0.932-1.852) | 0.119 | 1.299 (0.908-1.859) | 0.153 |
| Chronic Viral Hepatitis | 1.193 (0.482-2.954) | 0.703 | 1.336 (0.537-3.321) | 0.534 | 1.326 (0.533-3.300) | 0.544 | 1.501 (0.600-3.755) | 0.386 |
| Liver Disease | 1.191 (0.708-2.004) | 0.509 | 1.184 (0.698-2.009) | 0.531 | 1.276 (0.750-2.170) | 0.369 | 1.268 (0.739-2.173) | 0.389 |
| Acute Kidney Injury | 0.878 (0.656-1.175) | 0.381 | 0.823 (0.603-1.123) | 0.219 | 0.935 (0.690-1.266) | 0.664 | 0.887 (0.643-1.223) | 0.464 |
| **Labs** |  |  |  |  |  |  |  |  |
| WBC | 1.012 (0.995-1.028) | 0.162 | 1.018 (1.000-1.036) | 0.047 | 1.005 (0.987-1.023) | 0.605 | 1.010 (0.991-1.029) | 0.315 |
| Creatinine > 1.2 | 1.353 (1.108-1.654) | 0.003 | 1.337 (1.089-1.641) | 0.006 | 1.281 (1.042-1.575) | 0.019 | 1.288 (1.043-1.591) | 0.019 |
| Anion Gap | 1.042 (1.022-1.062) | <0.001 | 1.040 (1.019-1.061) | <0.001 | 1.032 (1.010-1.054) | 0.004 | 1.028 (1.005-1.051) | 0.015 |
| Potassium | 1.125 (1.019-1.241) | 0.019 | 1.103 (0.996-1.221) | 0.060 | 1.103 (0.994-1.225) | 0.064 | 1.081 (0.971-1.204) | 0.154 |
| ALT | 1.000 (0.999-1.001) | 0.727 | 1.000 (0.999-1.001) | 0.757 | 1.000 (0.999-1.001) | 0.688 | 1.000 (0.999-1.001) | 0.758 |
| **Hospital Conditions** |  |  |  |  |  |  |  |  |
| ICU | 1.350 (1.094-1.667) | 0.005 | 1.425 (1.149-1.768) | 0.001 | 1.382 (1.109-1.723) | 0.004 | 1.437 (1.148-1.800) | 0.002 |

Abbreviations: ACE, angiotensin-converting enzyme; ARBs, angiotensin II receptor blockers; BMI, body mass index; BP, blood pressure; SBP, systolic blood pressure; DBP, diastolic blood pressure; COPD, chronic obstructive pulmonary disease; HIV, human immunodeficient virus; WBC, white blood cell count; ALT, alanine aminotransferase

### Inpatient α_1_-blocker use and COVID-19 in-hospital mortality

We further assessed the impact of α_1_-blockers on mortality for patients with documented administration of α_1_-blockers while admitted to the hospital (N=343) compared to unexposed patients. We observed that inpatient α_1_-blocker use significantly reduced the risk of in-hospital mortality overall (OR 0.633; 95% CI 0.434-0.921; P=0.017) and among those hospitalized for at least 24 hours (OR 0.607; 95% CI, 0.415-0.890; P=0.01) (Table 2).

The survival analysis also showed improved in-hospital survival for those with inpatient α_1_-blocker use overall (HR 0.721; 95% CI, 0.572-0.908; P=0.006), and among patients hospitalized for at least 24 hours (HR 0.751; 95% CI, 0.593-0.950; P=0.017) (Table 3).

### Sensitivity analysis

We assessed the impact of potential unmeasured or uncontrolled confounding on the observed associations between inpatient α_1_-blockers use and COVID-19 in-hospital mortality using the E-value metric (Table S2).^16^ Based on this analysis, an unmeasured confounder would have to have an odds ratio of at least 1.25 (in the logistic regression analysis) or a hazard ratio of at least 1.34 (in the survival analysis) to cause the 95% confidence interval for the treatment effect of inpatient α_1_-blockers use to include the null.

### Age-stratified associations of α_1_-blockers and COVID-19 in-hospital mortality

We assessed the interaction between age and inpatient α_1_-blocker use for overall cohort and patients with hospital stay > 1 day, respectively, and we did not find any significance between the interaction and in-hospital mortality. Therefore, to identify differences in the treatment effect of α_1_-blockers on different age groups, we segmented the population into three age groups (45-65; 55-75; 65-89) and analyzed each group separately using logistic regression, adjusting for the same covariates as the unstratified analysis. The age groups were overlapped by 10 years to preserve sample size. Inpatient α_1_-blocker use was associated with a significantly lower risk of in-hospital mortality in the 45-65 (OR 0.384; 95% CI 0.164-0.896; P=0.027) and 55-75 age groups (OR 0.511; 95% CI 0.297-0.880; P=0.015), but not the 65-89 age group (OR 0.810; 95% CI 0.509-1.289; P=0.374) (Table 4).

**Table 4.** Age-stratified multivariable logistic regression and propensity-weighted logistic regression analyses for COVID-19 in-hospital mortality, by cohort (overall vs. hospitalization >1 day) and alpha-1-blocker exposure (any vs. inpatient use)

|  | Age 45-65 (N=1159) |  | Age 55-75(N=1312) |  | Age 65-89 (N=1176) |  |
| --- | --- | --- | --- | --- | --- | --- |
|  | Adjusted OR (95% CI) | p-value | Adjusted OR (95% CI) | p-value | Adjusted OR (95% CI) | p-value |
| **Multivariate logistic regression** |  |  |  |  |  |  |
| Overall cohort, any exposure | 0.511 (0.232-1.126) | 0.096 | 0.603 (0.362-1.003) | 0.051 | 0.908 (0.588-1.402) | 0.663 |
| Overall cohort, inpatient use | 0.384 (0.164-0.896) | 0.027 | 0.511 (0.297-0.880) | 0.015 | 0.810 (0.509-1.289) | 0.374 |
| Hospital stay > 1 day, any exposure | 0.530 (0.240-1.174) | 0.117 | 0.590 (0.352-0.987) | 0.045 | 0.880 (0.563-1.374) | 0.574 |
| Hospital stay > 1 day, inpatient use | 0.397 (0.170-0.927) | 0.033 | 0.507 (0.294-0.874) | 0.015 | 0.789 (0.492-1.265) | 0.325 |
| **Inverse propensity-weighted logistic regression** |  |  |  |  |  |  |
| Overall cohort, any exposure | 0.298 (0.132-0.672) | 0.004 | 0.552 (0.337-0.903) | 0.018 | 0.876 (0.557-1.379) | 0.568 |
| Overall cohort, inpatient use | 0.268 (0.109-0.658) | 0.004 | 0.424 (0.247-0.725) | 0.002 | 0.801 (0.487-1.317) | 0.382 |
| Hospital stay > 1 day, any exposure | 0.314 (0.141-0.698) | 0.005 | 0.535 (0.321-0.89) | 0.016 | 0.800 (0.500-1.280) | 0.353 |
| Hospital stay > 1 day, inpatient use | 0.262 (0.112-0.61) | 0.002 | 0.451 (0.268-0.759) | 0.003 | 0.752 (0.455-1.242) | 0.265 |

### Propensity score analysis

We further assessed the effect of α_1_-blockers on in-hospital mortality using propensity-weighted logistic regression. These yielded similar results as the multivariable regression and age-stratified analyses. Inpatient α_1_-blocker use was associated with reduced in-hospital mortality (OR 0.630; 95% CI 0.424-0.935; P=0.022) whereas α_1_-blocker exposure was not (OR 0.740; 95% CI 0.512-1.070; P=0.110) (Table S3). In age-stratified analyses, patients in younger age groups had greater benefit from α_1_-blocker exposure and inpatient α_1_-blocker use than patients in the oldest age group (Table 4).

## Discussion

Using a racially and ethnically diverse cohort from New York City comprising 2,627 men aged 45 or older hospitalized COVID-19 patients seen between February 24 and March 31, 2020, we found that inpatient use of α_1_-blockers was significantly associated with reduced in-hospital mortality, whereas outpatient exposures were not associated with benefit, although there was a trend towards significance. In age stratification analysis, we found that α_1_-blocker use was more beneficial to younger age groups.

Drug repurposing is the process of finding new indications for drugs already in clinical use. The appeal of rapidly validating and deploying an existing drug against a deadly global pandemic is clear, especially if the drug is widely available and affordable. Dexamethasone, now a standard in COVID-19 treatment, is an example of a commonly used drug repurposed for a new indication.^3^ However, the saga of hydroxychloroquine, which was touted as a cure early in the pandemic but has since proven ineffective, is a cautionary tale.^17^ The allure of rapid drug repurposing must be balanced against rigorous scientific method.

α_1_-blockers, commonly used to treat benign prostatic hyperplasia and hypertension, have become a target for drug repurposing due to pre-clinical data linking α_1_-adrenergic signaling to pro-inflammatory cytokines which may contribute to dysregulated immunity and adverse outcomes in COVID-19.^1,2,11^ These pre-clinical findings have been bolstered by recent retrospective clinical analyses linking α_1_-blockers with improved outcomes in hospitalized patients with both COVID-19 and non-COVID-19 respiratory infections.^12,13^ In a large COVID-19 cohort drawn from the US Veterans Health Administration hospital system, outpatient α_1_-blocker exposure was associated with a relative risk reduction of 18% for in-hospital mortality compared to matched controls.^13^ Interestingly, the non-selective α_1_-blocker doxazosin, which inhibits all three α_1_-adrenergic receptor subtypes (α_1A_, α_1B,_ α_1D_), was associated with a greater relative risk reduction (74%) than the uroselective (α_1A_, α_1D_) α_1_-blocker tamsulosin (18%).

In the present study, we found that in-hospital use of α_1_-blockers was consistently and independently associated with reduced in-hospital mortality using both multivariable regression and propensity score-based methods. In age-stratified analyses, we observed that this protective effect was more pronounced in relatively younger age groups. In contrast to the studies by Koenecke et al. and Rose et al., which defined α_1_-blocker exposure based on outpatient prescriptions only, we were able to use inpatient medication administration records to identify patients treated with α_1_-blockers during their COVID-19 hospitalization. These results lend additional support the hypothesis that α_1_-blockers may have a beneficial effect in COVID-19.

Our results also include tests of association between other common medication classes and COVID-19 outcomes, including beta blockers, angiotensin-converting enzyme (ACEi) inhibitors, angiotensin II receptor blockers (ARBs), and glucocorticoids. Results pertaining to these other medications should be taken in the context of a selected cohort designed to study α_1_-blocker use and COVID-19 outcomes. Exposures to these other classes were not ascertained with the same granularity as they were for the primary exposure. That said, it is interesting to note that α_1_-blocker and beta blocker exposure were associated with opposite COVID-19 outcomes in our cohort. There is evidence to suggest that β-adrenergic signaling can promote an anti-inflammatory M2 phenotype in macrophages, in contrast to the pro-inflammatory effect of α_1_-adrenergic signaling.^11,18^ Additional efforts to dissect the interactions between adrenergic signaling and the COVID-19 immune response are warranted. A prior diagnosis of asthma was associated with reduced in-hospital mortality in this analysis. While this observation deserves further scrutiny, it is conceivable that early exposure to inhaled glucocorticoids or β-adrenergic agonists may have contributed to this signal.

### Limitations

Our study has several limitations. The cohort did not include women since most α_1_-blockers were prescribed to men, most likely for benign prostatic hyperplasia. Male sex is a recognized risk factor for adverse COVID-19 outcomes, possibly due to sex-specific differences in immunity.^19^ Thus, these results may not extrapolate to women. We did not account for different types of α_1_-blockers, which differentially target the three α_1_-adrenergic receptor subtypes. Importantly, a causal relationship cannot be definitively established between α_1_-blockers and improved COVID-19 outcomes in this retrospective study. Several confounders, such as older age and comorbidities were more common in the α_1_-blocker group. However, these adverse risk factors would be expected to bias the study result towards the null rather than inflate a protective association. Furthermore, our findings are consistent with prior data, and were robust to different methods of adjustment for confounding and sensitivity analysis. The ongoing randomized clinical trial of prazosin against placebo among hospitalized COVID-19 patients (NCT04365257) will include women and provide more definitive data on the therapeutic value of α_1_-blockers. Finally, our study is based on a single center EMR and medication via outpatient use cannot be tracked for adherence. Therefore, analyzing in-hospital medication administration is more robust. Furthermore, EMR in MSHS is the one of the largest and most comprehensive EMR systems, representing racial/ethnic diversity in New York City, and EMR implementation is from various data sources, so our findings are supported by large patient cohort and were robust from applying multiple methods of adjustment for confounding.

## Conclusions

In conclusion, this retrospective study found a protective association between α_1_-blockers use and COVID-19 outcomes in a cohort of hospitalized men. These results augment the rationale for studying and repurposing α_1_-blockers as a COVID-19 therapeutic. We await the results of the ongoing randomized clinical trial.

## Data Availability

The Mount Sinai Health System database is not publicly available. We utilized Python and R, and their open-source libraries to conduct our analysis tailored to MSH data. Therefore, we are unable to publicly release the code since it is useless without the dataset available.

## Acknowledgement

The authors would like to acknowledge Sema4 IT for computational support hosted on AWS. JV is supported by grants from Microsoft Research and FastGrants.

## Author contributions

conceived and designed the study: WKO, LL and SL; performed data extraction from MSHS: SL, ZW, ES; performed statistical analysis: SL; contributed clinical interpretation: TJ, MFK, CB, JV, NP, EES, RP, RC, LL, WKO; wrote and edited the paper: SL, TJ, MFK, CB, JV, NP, EES, RP, RC, LL, WKO.

## Conflict of interest

Sema4 is a for-profit company currently majority owned by the Icahn School of Medicine at Mount Sinai (ISMMS). RC, EES, LL and WKO receive compensation from Sema4 that includes equity in the company. In addition to their roles with Sema4, RC, EES, LL and WKO remain affiliated with ISSMS as part-time employees and faculty members. WKO also has consulted for AAA, Astellas, AstraZeneca, Bayer, Conjupro, Foundry, Janssen, Merck, Sanofi and TeneoBio. The JHU filed a patent application on the use of various drugs to prevent cytokine release syndromes, on which NP, BV, KWK, and SZ are listed as inventors. JHU will not assert patent rights from this filing for treatment related to COVID-19. NP is a founder of, consultant to and holds equity in Thrive an Exact company. NP is a founder of and holds equity in Personal Genome Diagnostics. NP is an advisor to and holds equity in Cage Pharma, ManaTbio and NeoPhore. CB is a consultant to Depuy-Synthes and Bionaut Labs. CB, MFK, BV, KWK, and NP are also inventors on technologies unrelated or indirectly related to the work described in this article. Licenses to these technologies are or will be associated with equity or royalty payments to the inventors, as well as to JHU. The terms of all these arrangements are being managed by JHU in accordance with its conflict of interest policies.

## SUPPLEMENTARY MATERIALS

**Figure S1.**
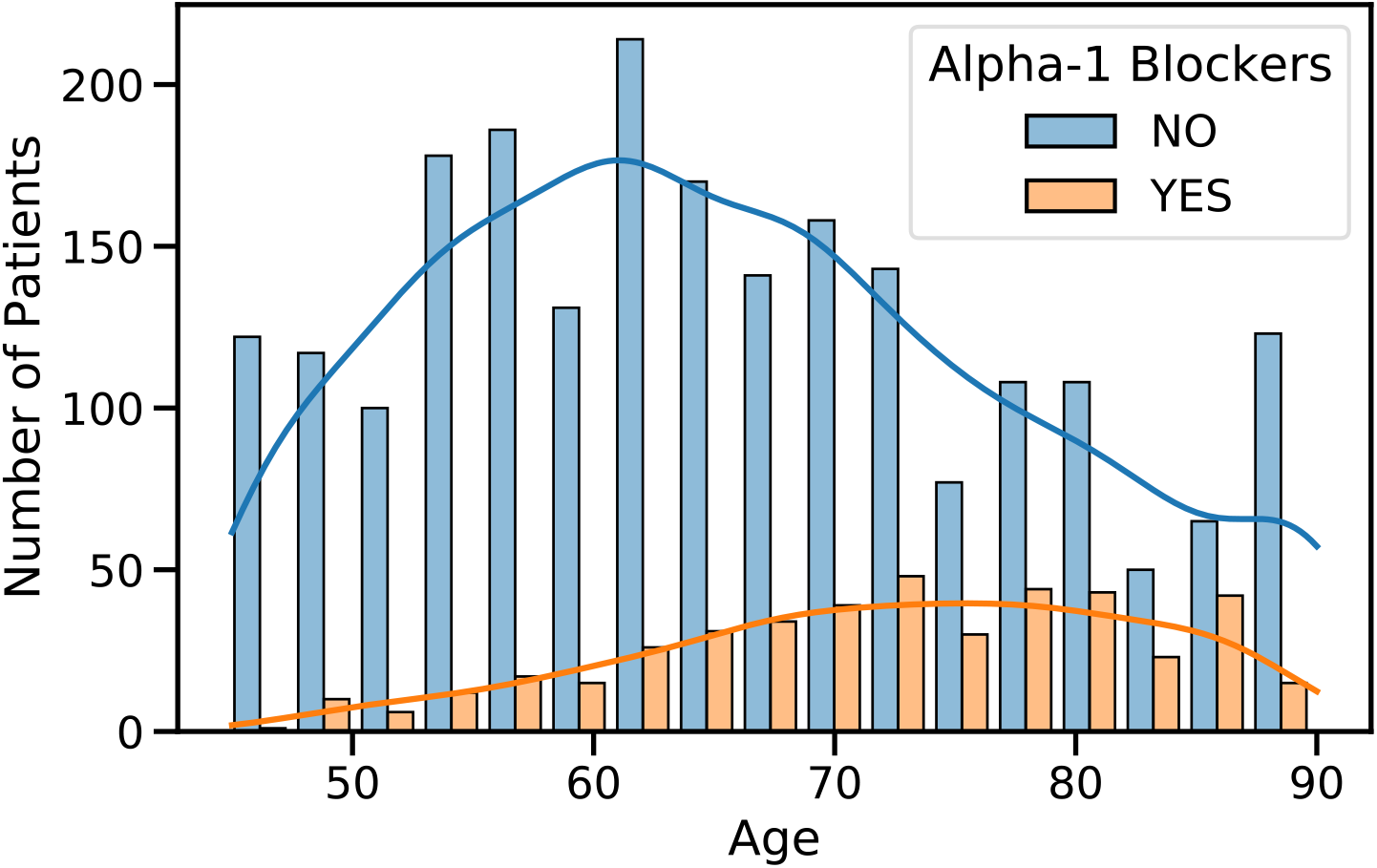
Age distributions of α_1_-blockers exposed and unexposed groups.

**Table S1a.** Medication categories and individual medications.

| Medication Category | Medication Names |
| --- | --- |
| Alpha-1 Blockers | Tamsulosin, Alfuzosin, Silodosin, Terazosin, Doxazosin, Prazosin |
| ACE inhibitors | Benazepril hydrochloride, Captopril, Enalapril maleate, Fosinopril sodium, Lisinopril, Moexipril, Perindopril, Quinapril hydrochloride, Ramipril, Trandolapril |
| ARBs | Azilsartan, Candesartan, Eprosartan mesylate, Irbesartan, losartan potassium, Olmesartan, Telmisartan, Valsartan |
| Diuretics | Chlorthalidone, Chlorothiazide, Hydrochlorothiazide, Indapamide, Metolazone, Amiloride hydrochloride, Spironolactone, Triamterene, Eplerenone, Furosemide, Bumetanide, Torsemide, Ethacrynic acid |
| Beta Blockers | Acebutolol, Atenolol, Betaxolol, Bisoprolol fumarate, Carteolol hydrochloride, Metoprolol, Nadolol, Nebivolol, Penbutolol sulfate, Pindolol, Propranolol hydrochloride, Solotol hydrochloride, Timolol maleate |
| Calcium-channel Blockers | Amlodipine, Bepridil, Diltiazem hydrochloride, Felodipine, Isradipine, Nicardipine, Nifedipine, Nisoldipine, Verapamil hydrochloride |
| Statin | Atorvastatin, Fluvastatin, Lovastatin, Pitavastatin, Pravastatin, Rosuvastatin, Simvastatin |
| Glucocorticoid | Methylprednisolone, Prednisone, Hydrocortisone, Dexamethasone, Fludrocortisone, Triamcinolone, Prednisolone, Betamethasone |

**Table S1b.** Number of patients exposed to glucocorticoid medications.

| Drug Name | Number of Patients |
| --- | --- |
| Methylprednisolone | 621 |
| Prednisone | 269 |
| Hydrocortisone | 242 |
| Dexamethasone | 189 |
| Fludrocortisone | 41 |
| Triamcinolone | 29 |
| Prednisolone | 23 |
| Betamethasone | 10 |

**Table S2.** Sensitivity analysis using E-values to assess the threshold effect size of unmeasured confounders required to nullify the significant associations between inpatient alpha-1-blocker use and COVID-19 in-hospital mortality.

|  | Point estimate for OR<br>(Upper limit of CI) | Point estimate for HR<br>(Upper limit of CI) |
| --- | --- | --- |
| Overall cohort, inpatient use | 1.83 (1.25) | 1.82 (1.34) |
| Hospital stay > 1 day, inpatient use | 1.89 (1.31) | 1.74 (1.23) |

**Table S3.** Inverse propensity-weighted logistic regression for COVID-19 in-hospital mortality, by cohort (overall vs. hospitalization >1 day) and alpha-1-blocker exposure (any vs. inpatient use)

|  | Adjusted OR (95% CI) | p-value |
| --- | --- | --- |
| Overall cohort, any exposure | 0.740 (0.512-1.070) | 0.110 |
| Overall cohort, inpatient use | 0.630 (0.424-0.935) | 0.022 |
| Hospital stay > 1 day, any exposure | 0.760 (0.520-1.110) | 0.157 |
| Hospital stay > 1 day, inpatient use | 0.622 (0.419-0.924) | 0.019 |

## Notes

### Author Declarations

This study was approved by the Mount Sinai institutional review board (IRB): IRB-17-01245

## References

1. Qin C, Zhou L, Hu Z, et al. Dysregulation of Immune Response in Patients With Coronavirus 2019 (COVID-19) in Wuhan, China. Clinical Infectious Diseases. 2020;71(15):762–768. doi:10.1093/cid/ciaa248

2. Del Valle DM, Kim-Schulze S, Huang H-H, et al. An inflammatory cytokine signature predicts COVID-19 severity and survival. Nature Medicine. 2020;26(10):1636–1643. doi:10.1038/s41591-020-1051-9

3. The RECOVERY Collaborative Group. Dexamethasone in Hospitalized Patients with Covid-19 — Preliminary Report. New England Journal of Medicine. 2020;0(0):null. doi:10.1056/NEJMoa2021436

4. Salama C, Han J, Yau L, et al. Tocilizumab in Patients Hospitalized with Covid-19 Pneumonia. New England Journal of Medicine. 2021;384(1):20–30. doi:10.1056/NEJMoa2030340

5. Hermine O, Mariette X, Tharaux P-L, et al. Effect of Tocilizumab vs Usual Care in Adults Hospitalized With COVID-19 and Moderate or Severe Pneumonia: A Randomized Clinical Trial. JAMA Intern Med. 2021;181(1):32. doi:10.1001/jamainternmed.2020.6820

6. The REMAP-CAP Investigators, Gordon AC, Mouncey PR, et al. Interleukin-6 Receptor Antagonists in Critically Ill Patients with Covid-19 – Preliminary report. medRxiv. Published online January 7, 2021:2021.01.07.21249390. doi:10.1101/2021.01.07.21249390

7. RECOVERY Collaborative Group, Horby PW, Pessoa-Amorim G, et al. Tocilizumab in patients admitted to hospital with COVID-19 (RECOVERY): preliminary results of a randomised, controlled, open-label, platform trial. medRxiv. Published online February 11, 2021:2021.02.11.21249258. doi:10.1101/2021.02.11.21249258

8. Benjamin D. Randomized Master Protocol for Immune Modulators for Treating COVID-19. clinicaltrials.gov; 2021. Accessed February 15, 2021. https://clinicaltrials.gov/ct2/show/NCT04593940

9. Tardif J-C, Bouabdallaoui N, L’Allier PL, et al. Efficacy of Colchicine in Non-Hospitalized Patients with COVID-19. medRxiv. Published online January 27, 2021:2021.01.26.21250494. doi:10.1101/2021.01.26.21250494

10. Kalil AC, Patterson TF, Mehta AK, et al. Baricitinib plus Remdesivir for Hospitalized Adults with Covid-19. New England Journal of Medicine. 2020;0(0):null. doi:10.1056/NEJMoa2031994

11. Staedtke V, Bai R-Y, Kim K, et al. Disruption of a self-amplifying catecholamine loop reduces cytokine release syndrome. Nature. 2018;564(7735):273–277. doi:10.1038/s41586-018-0774-y

12. Koenecke A, Powell M, Xiong R, et al. Alpha-1 adrenergic receptor antagonists to prevent hyperinflammation and death from lower respiratory tract infection. arXiv:200410117 [q-bio]. Published online September 8, 2020. Accessed February 16, 2021. http://arxiv.org/abs/2004.10117

13. Rose L, Graham L, Koenecke A, et al. The Association Between Alpha-1 Adrenergic Receptor Antagonists and In-Hospital Mortality from COVID-19. medRxiv. Published online February 11, 2021:2020.12.18.20248346. doi:10.1101/2020.12.18.20248346

14. Konig MF, Powell M, Staedtke V, et al. Preventing cytokine storm syndrome in COVID-19 using α-1 adrenergic receptor antagonists. J Clin Invest. 2020;130(7):3345–3347. doi:10.1172/JCI139642

15. Demidenko E. Sample size determination for logistic regression revisited. Stat Med. 2007;26(18):3385–3397. doi:10.1002/sim.2771

16. VanderWeele TJ, Ding P. Sensitivity Analysis in Observational Research: Introducing the E-Value. Ann Intern Med. 2017;167(4):268–274. doi:10.7326/M16-2607

17. Singh B, Ryan H, Kredo T, Chaplin M, Fletcher T. Chloroquine or hydroxychloroquine for prevention and treatment of COVID-19. Cochrane Database of Systematic Reviews. 2021;(2). doi:10.1002/14651858.CD013587.pub2

18. Lamkin DM, Ho H-Y, Ong TH, et al. β-adrenergic-stimulated macrophages: Comprehensive localization in the M1–M2 spectrum. Brain Behav Immun. 2016;57:338–346. doi:10.1016/j.bbi.2016.07.162

19. Takahashi T, Iwasaki A. Sex differences in immune responses. Science. 2021;371(6527):347–348. doi:10.1126/science.abe7199

